# Estimating the epidemic growth dynamics within the first week

**DOI:** 10.1101/2020.08.14.20170878

**Authors:** Enzo Fioriti, Marta Chinnici, Andrea Arbore, Nicola Sigismondi, Ivan Roselli

**Affiliations:** C.R. ENEA Casaccia; Srl Ict Technical consultant

## Abstract

Information about the early growth of infectious outbreaks are indispensable to estimate the epidemic spreading. A large number of mathematical tools have been developed to this end, facing as much large number of different dynamic evolutions, ranging from sub-linear to super-exponential growth. Of course, the crucial point is that we do not have enough data during the initial outbreak phase to make reliable inferences. Here we propose a methodology to estimate the epidemic growth dynamics from the infected cumulative data of just a week, provided a surveillance system is available over the whole territory. The methodology, based on the Newcomb-Benford Law, is applied to Italian covid 19 case-study. Results show that it is possible to discriminate the epidemic dynamics using the first seven data points collected over fifty Italian cities. Moreover, the form of the most probable approximating function of the growth, within a six weeks epidemic scenario, is identified.

## Introduction

During an outbreak, a major issue to stop or mitigate the virus diffusion is gathering information as soon as possible about the nature of the epidemic from a mathematical point of view. Pandemic outbreaks allow a reasonable amount of data to be available only *ex-post* the event, therefore the analysis is severely limited. On the other hand, well-timed information about the epidemic growth is extremely precious, and justify any effort in this direction. A panoply of tools are available, but their accuracies are subject to limitations, due to the small number of data points available, which also restricts the choice of models for the epidemic curve that is a time-series of the cumulative number of cases per day [1, 2]. These curves are produced by different dynamics, ranging from sub-linear to super-exponential, giving rise to a diversity of the early growth profiles that has deep implications for the estimation of the disease transmission and for the countermeasures implementation [3]. Therefore, a fast detection and estimation at least of the basic outbreak characteristics, would be extremely useful, but the mathematical tools able to deal with those very few data are rare. Moreover, epidemic data gathered on the field are always polluted by human errors, different collection methods, limited territorial coverage, irregular or random sampling. Even the most recent sophisticated signal processing techniques such as the Graph spectral analysis, the Compressive Sensing, the Signal on Graphs method are affected by these problems [4 – 21]. However, the Newcomb-Benford law (NBL) seems able to reveal the epidemic dynamics using only a week of infection data. NBL is the statistic of the first (actually also of the second, third and so on) digit for a set of numbers, discovered and rediscovered independently by S. Newcomb and F. Benford, today known as the Benford law [22, 23, 24]. Its wide popularity is due to the apparent ubiquity of the Benford distribution and to the extreme simplicity of the calculation procedure involved. In recent years, the NBL has been used to discover fiscal frauds and to confirm scientific data reliability, including epidemic data [25]. In our work, we are interested to study the capabilities of the NBL to predict the outbreak growth dynamics using very few initial data. It is only necessary to collect the cumulative data of the daily infected over a week in some of the most important cities involved in the outbreak, to form a unique sequence of these numbers and then to calculate the first digit distribution. Given the limited amount of data points, the calculated and the actual Benford distribution will not coincide exactly, thus to characterize the accuracy an appropriate goodness-of-fit (gof) parameter is used. At this point, by trial-and-error or by any numerical technique, an approximating function is chosen: if its first digit distribution is congruent with that of the Italian cities, we can consider the approximating function as an accurate approximation to the real cumulative curve. In the following sections we will show how this is possible invoking the Theorems of Berger & Hill and the ergodicity of the epidemic SIS process during the initial expansion phase [26].

### The Newcomb – Benford Law

Before presenting the essential points of our proposal, we have to introduce the main statistical tool, namely the Newcomb-Benford Law. Since an extensive treatment can be found in the fundamental work of Hill and Berger [22, 23, 24], we will give just a brief introduction to the NBL from a practical point of view. S. Newcomb and F. Benford independently observed that the leading digits in many real-life numerical data sets such as macroeconomic, census, financial, fiscal data, were not distributed uniformly, as the common sense would suggest, instead they follow the logarithmic distribution of Figure 1a:

**Figure 1.**
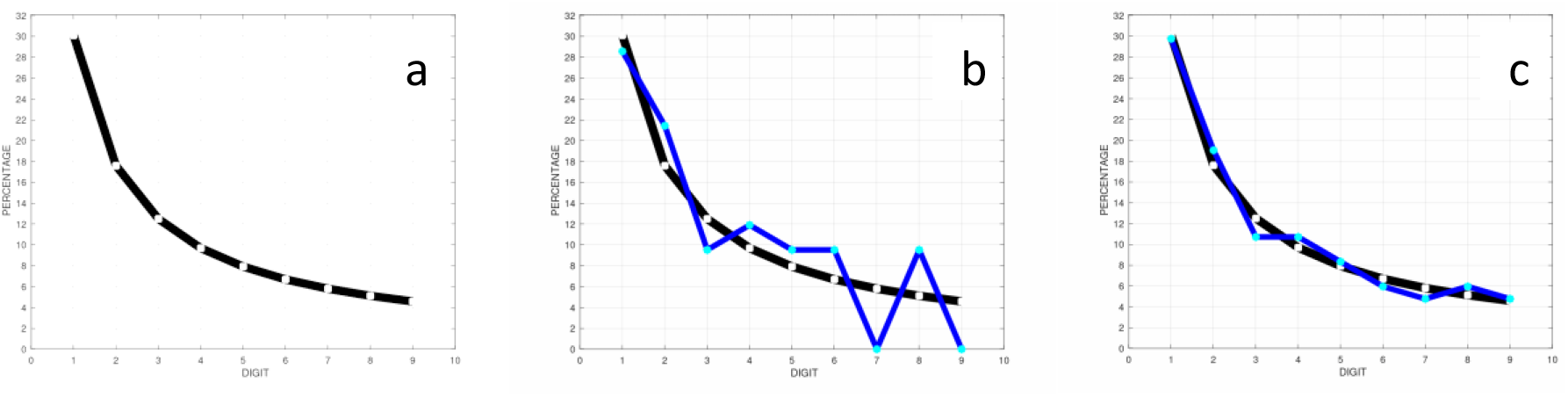
a) The Benford distribution: digits 1 – 9 and their percentages: 30.1, 17.6, 12.5, 9.69, 7.92, 6.69 5.80, 5.12, 4.58. First digit distribution with a limited amount of data: b) 2*^n^* first 42 data-points (blue) and c) 2*^n^* first 84 data-points (blue). The curve of Figure 1c is close to the real Benford distribution; in this case *gof* = 0.99813, to be compared to the *gof* = 3.5803 of the previous one.

**Figure 2.**
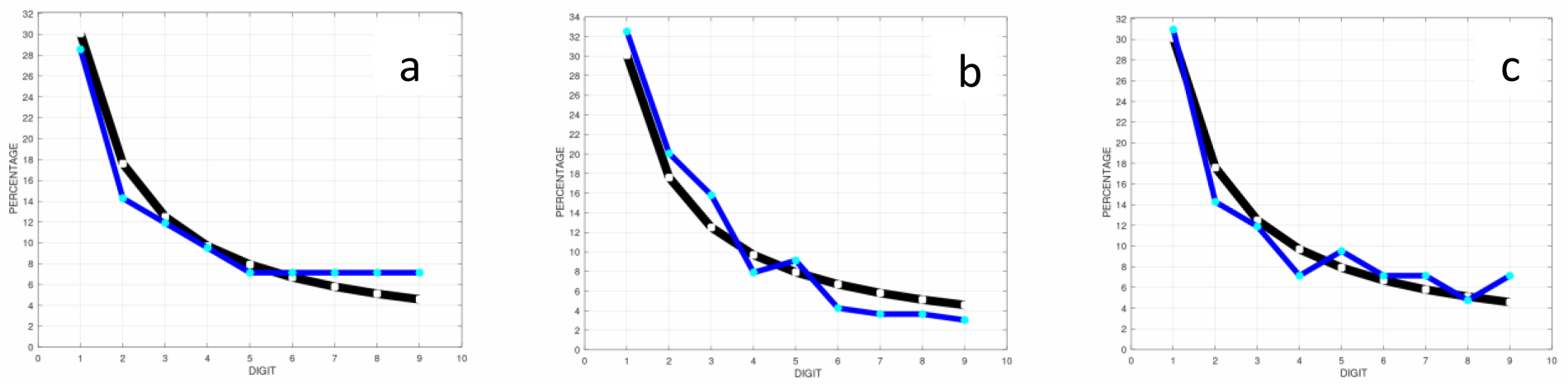
Visually all the blue curves are similar to the Benford distribution (black). a) in blue: IT_*real*, the Italian cumulative data first digit distribution, *gof* = 1.7313. b) blue: the 50_*cities* first digit distribution, *gof*= 1.8081. c) blue: the logistic curve, *gof* = 1.8206.

More formally, the Benford distribution is a logarithmic curve:

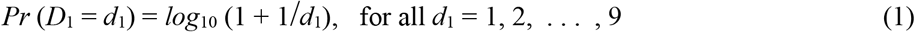

where *D*_1_ is the first *significant* decimal digit. First significant digit means 4 in 0.467, 5 in 58.34, 3.7 in 3.7*10^2^, 8.21 in 8.21*10^-4^. When a set of numbers follows exactly the NBL, the digit “1” will appear about the 30% of the times, the digit “2” the 17%, “3” the 12% etc.. The Generalized NB Law considers also the second, third etc. digit, but here we are interested only to the first one.

The compliance to the Benford distribution may be found also in some natural data-sets, such as molecular weight tables, sport statistics, drainage areas of rivers, that taken individually do not follow the NBL: what satisfies completely the NBL is the union of all those data-sets.

Some numerical sequences follows strictly the Benford first digit distribution. Let us consider the sequence of powers 2*^n^*: {2 4 8 16 32 64 128 256 512 1024 2048 4096 8192 16384 32768 65536 131072 262144 524288 1048576 …}. The powers of 2 follow the Benford distribution (we say “is Benford”, for short), but to verify it numerically, we would need of a large set of numbers. This constitutes one first difficulty, because it is not easy to determine the minimum cardinality of the set that guarantees *a priori* to reveal exactly the Benford distribution. Moreover, most of the time we do not have enough data to satisfy correctly the NBL, and, as a consequence, an error is introduced. In Figure 1b, 1c the effect of a limited data set is clearly illustrated: using only 42 data points we obtain a poor fit to the actual Benford distribution, although simply doubling the data-points reduces greatly the error. Hence, it is convenient to use a goodness-of-fit parameter to estimate the error committed; here, to this end, we consider the standard measure:

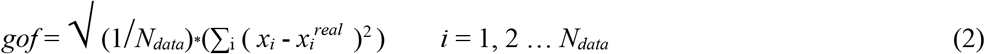

but other statistical tests may be used as well. Of course, generally the first digit distribution of a data-set may or may not be a Benford (first digit) distribution; if this is the case, it will be clearly specified. In any case, the gof indicates the distance between the calculated firs digit distribution and the Benford distribution.

### Theoretical justification and technical background

The main idea is to estimate an approximating function for the epidemic growth curve within a time horizon of *T_f_* days, using only the first seven epidemic data points of fifty Italian cities, accounting for about the 30% of the population, considered as a unique sequence formed of 50×7 data-points, called 50_*cities* sequence. We show that the first digit distribution of the 50_*cities* sequence converges to the first digit distribution of the cumulative daily infected Italian national sequence, formed summing the infected over all the national territory each day during the first *T_f_* days of the epidemic ascending phase. Therefore, if the convergence exists, it is possible to know the compliance to Benford for the cumulative Italian curve in advance of *T_f_ –* 7 days. In turn, the level of compliance is used as a criterion to predict the accuracy of an approximating function to the real epidemic national curve during the initial phase of *T_f_* days (starting from the 21 *th* February 2020).

Now we will sketch the theoretical justification of the above method. First of all, we have to make sure that the Italian national cumulative data first digit distribution is Benford during the initial spreading period. Based on the Berger & Hill theorems [22, 23], if the sequence is a power law, exponential or super-exponential, its first digit distribution is almost always Benford. Thus, we have only a sufficient condition. Below some of the main results of Berger & Hill to support our method (formal demonstrations can be found in [23, 24, 25]).

Theorem (Berger & Hill 1). *None of the classical probability distributions or random variables, such as the normal, uniform, exponential, beta, binomial, or gamma distributions are Benford. Specifically, no uniform distribution is even close to Benford, no matter how large its range or how it is centered. However, some distributions come close to being Benford, such as the Pareto and the Log-normal distribution*.

The next Theorem states that the solutions of an ordinary or differential equation system such as many of the classical epidemic models, under general conditions are Benford. Thus, the NBL is not restricted to discrete dynamics; on the other hand, general results for partial differential, delay or integro-differential equations are not known.

Theorem (Berger & Hill 2). *Consider the dynamic system*:

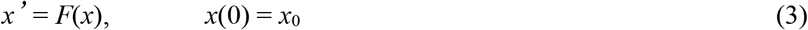

*where F*: ℝ → ℝ *is continuously differentiable with F*(0) = 0, and *x*_0_ ∈ ℝ. *Let F*: ℝ → ℝ be *C*^2^ *with F*(0) = 0 *and assume F*’(0) < 0. *Then, for every x*_0_ ≠ 0 *sufficiently close to* 0, *the solution of the system is Benford*.

Related to Theorem 2 we have the so-called Shadowing Lemma. The Lemma describes the behavior of the pseudo-trajectories (sequences) near a locally structurally stable hyperbolic invariant set [22].

Shadowing Lemma. *Let T:* ℝ → ℝ *be a map, and β a real number with |β| >* 1. If 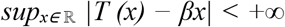 *then there exists, for every x ∈* ℝ, *one and only one point x° such that the sequence (T^n^(x)−β^n^x°) is bounded*

This means that every hyperbolic set has the shadowing property, thus every pseudo-trajectory or sequence stays uniformly close to some true trajectory, i.e. a pseudo-trajectory is “shadowed” by a true one. Considering the epidemic curve as the solution of a dynamic system, the Shadowing Lemma allows to believe that very close to it a pseudo-sequence exists, is related to the dynamic system and is structurally stable. In other words, finding an approximating function to the cumulative epidemic Italian curve would not be a mere accident, at least locally. Moreover, small perturbations of the system initial conditions do not change the approximation: therefore, errors during the initial data collections do not alter the result.

Theorem (Berger & Hill 3). *Let X be exponential with mean* 1, *that is F_X_(t)* = max(0, 1 – *e*^-^*^t^*), *t* ∈ ℝ. Even *though X is not exactly Benford, it is close to being Benford for all t* ∈ [1, 10).

Theorem (Berger & Hill 4). *The sequences:* 2*^n^*, 3*^n^ are Benford; n, n* +1, *n*!, √10*^n^*, 10*^n^*, 4_*_4*^n^ are not. In general, a^n^x^b^, with a >* 0 *and b >* 1 *is Benford almost always, but not always, therefore x*^2^ *is almost always Benford. Moreover, every mixture of* 2*^n^ with a random unbiased sequence, is Benford*.

Berger and Hill also state that apart from some particular cases, processes with linear growth are not Benford. This allows to identify the slow epidemic growths, that are a phenomenon more common than previously though [1]. By slow we mean a linear, sub-linear or a polynomial growth.

Theorem (Berger & Hill 5). *If X and Y are Benford sequences, also their sum X* + *Y is Benford. If the sequence Z is not Benford, X* + *Y* + *Z is Benford*.

Hence, if the cumulative infected sequence of the Italian cities are Benford, also the national cumulative sequence is Benford as well, during the weeks of the increasing phase. Note that an epidemic cumulative sequence cannot be random, being non-decreasing, therefore by Theorems Berger & Hill 1, 3, 4, 5 the fast cumulative epidemic curves are all Benford, but could exist also fast non-Benford curves in particular circumstances. Thus, we cannot rule out the possibility of non-Benford growth curves to have a fast dynamics, thought this would be seldom the case, see Table 1.

**Table 1.** – Various approximating functions are ordered according to the gof value (second column). In the last column is described the speed of growth within the first 42 days with respect to the real speed of growth of the case-study. The gof of the actual Italian cumulative curve *(IT*_*real)* is 1.7313, very close to the 50_*cities* gof; the best approximating function function is the cubic *I*(*t_i_*)*n*^3^. Samples is the number of data-points used to calculate the first digit distribution and therefore also the gof; they are 42, except for the 50_*cities* gof, whose distribution is calculated using not more than 294 data-points, the first 7 data from each city. In orange * are indicated the non-congruent dynamics: for example, 4*4*^n^* has a very large gof, nonetheless belongs to the class of fast dynamics, instead to that of the slow one.

| function | Benford goodness-of-fit |  | samples | growth to 42 days |
| --- | --- | --- | --- | --- |
| $5 \cdot 3^n$ | 0.9182 | | 42 | very fast |
| $12 \cdot (1.342)^n$ | 1.0269 | | 42 | fast |
| $3 \cdot 5^n$ | 1.4893 | | 42 | very fast |
| $16 \cdot n^3$ | 1.4934 | | 42 | fast |
| $2 \cdot 8^n$ | 1.5819 | | 42 | very fast |
| $2 \cdot 9^n$ | 1.6308 | | 42 | very fast |
| *IT_real* | **1.7313** | - | 42 | - |
| *50_cities* | **1.8081** | +4.4 % | 294 | - |
| $1.675 \cdot n^3$ | **1.8110** | +4.6 % | 42 | as *IT_real* |
| *logistic* | **1.8206** | +5.1 % | 42 | as *IT_real* |
| $2 \cdot 7^n$ | 2.0493 | +18.0 % | 42 | very fast * |
| $16 \cdot n^4$ | 3.1133 | | 42 | very fast * |
| $8 \cdot 2^n$ | 3.1237 | | 42 | very fast * |
| $16 \cdot n^2$ | 5.0668 | | 42 | slow |
| $3 \cdot 6^n$ | 5.6255 | | 42 | very fast * |
| $16 \cdot n$ | 6.9965 | | 42 | linear |
| $4 \cdot 4^n$ | 7.1002 | | 42 | very fast * |
| $16 \cdot \sqrt{n}$ | 10.783 | | 42 | Sub-linear |
| $50 \cdot \log_{10}(n)$ | 15.312 | | 42 | Sub-linear |

Now we can discuss the convergence of the 50_*cities* distribution to the distribution of the cumulative daily infected of the Italian national sequence after *T_f_* days. The 50_*cities* sequence is the union of the first 7 days data for each city, but to fix ideas, let us consider only three cities, A, B, C, whose sequences of seven elements are:

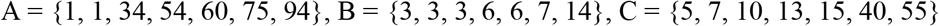

Thus the union *U* of A, B, C reads:

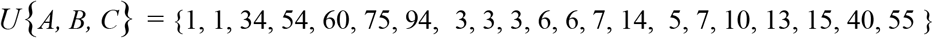

The cumulative number of cases per day is simply the total sum per day of the infected cases collected over all the *N_IT_* towns and cities of Italy, during the first *T_f_* days of the outbreak:

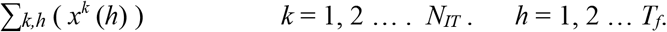

Indicating B(…) as the operator of the first digit distribution calculation, results:

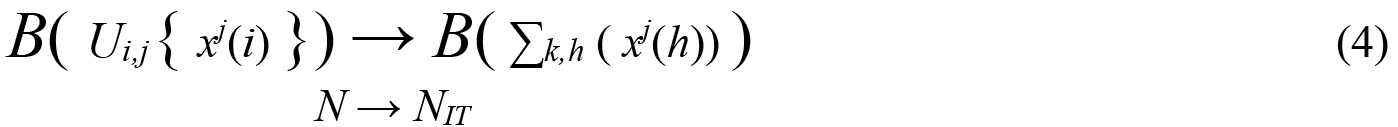

where *i =* 1, 2, *… m. j* = 1, 2, … *N*. *k* = 1, 2, … *N_IT_* . *h* = 1, 2, … *T_f_*.

In the trivial case, when the sequences *x^j^* are (almost all) Benford with *m* = *T_f_* and *N* = *N_IT_*, Berger & Hill 5 guarantees that the summation sequence is Benford. On the other hand, the union of the sequences is Benford too, thus the (4) is true.

When *m* = 7, since it is not known an analytical method to determine *a priori* the values of *m* that guarantees a small *gof*, one can only calculate the first digit distribution *B(U_i,j_{ x^j^(i)* }) and compare it to the Benford distribution of Figure 1a. If the *gof* stays high, the simplest heuristics would be to increase *m*, yet reducing the forecasting time-span of *T_f_ – m*. Of course, the number of city-sequences *N* may well be expanded to all the cities *N_IT_*: here we have restricted it to fifty cities only for demonstration purposes. Therefore, we do know that (4) is true, but cannot determine the minimum *m* necessary. Actually, we have chosen *m* = 7 because it is well below the prediction thresholds often suggested in the literature [2, 3]. Instead, to determine *T_f_*, we consider that during the initial phase of the outbreak if:

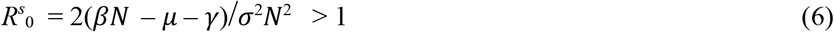

(where *μ* is the birth rate, *γ* is the cure rate, and *β* is the contact rate), the overall epidemic process is ergodic [26] . Hence, all the local realizations have similar statistics, are non random non decreasing sequences, and by the Shadowing lemma there exists an approximant function *f* to 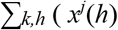:

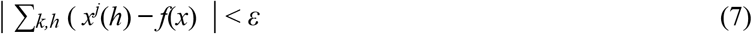

As a consequence, the first digit distribution of *f*(*x*) approximates that of 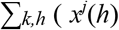, but by the (4) also that of *B*(*U_i,j_{ x^j^*(*i*) }). Therefore *T_f_* can be determined heuristically or numerically as the value that get *B(f(x))* closer to *B*(*U_i,j_{ x^j^*(*i*) }) in terms of Benford goodness-of-fit. From Table 1 it is readily seen that both the cubic and the logistic curve approximate the *IT_real* gof very well for *T_f_* = 42. In addition, *T_f_* = 42 is very close to the inflection point of the real Italian cumulative curve, which indicates the end of the initial phase for the outbreak.

### Application to the Italian case-study

Basically, we have three dynamics, very fast, fast, and slow (see Figure 3); we want to determine which one of them is prevailing by means of the Benford’ gof, and possibly to find an approximating function.

**Figure 3.**
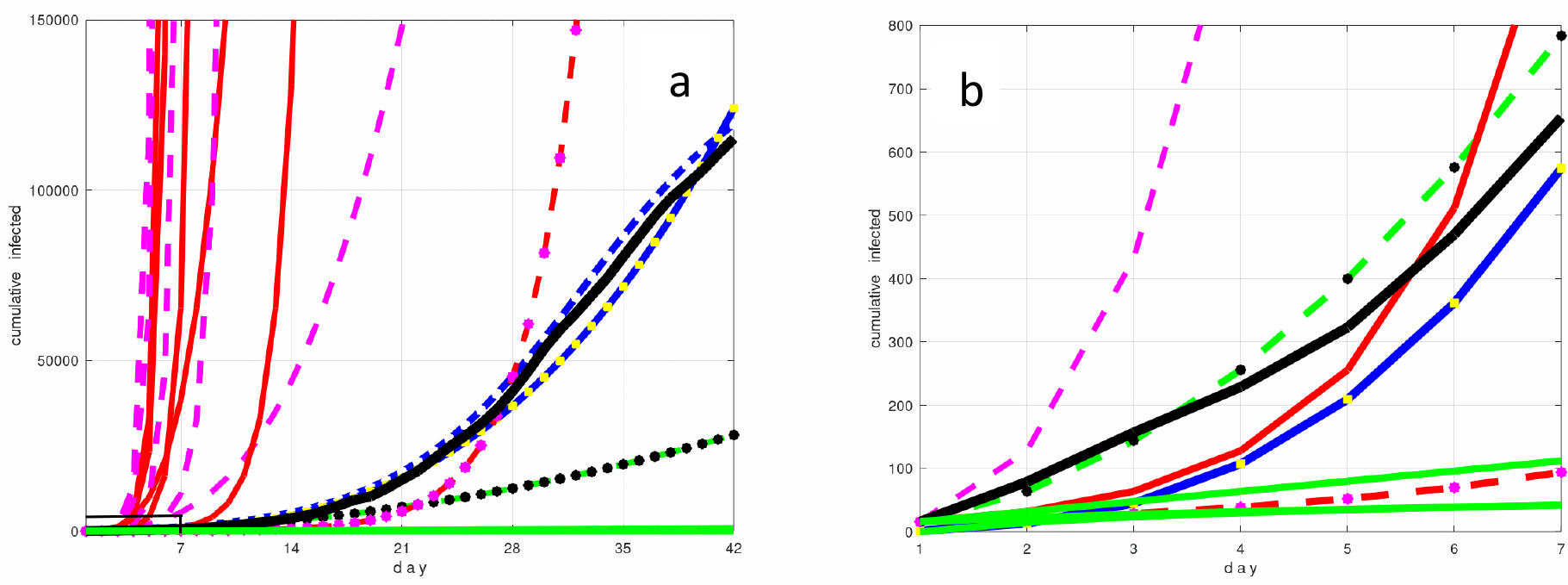
The first day is the 21 *th* February 2020. a) 42 days scenario. From left to right in the color code the dynamics of Table 1; dotted magenta and red: very fast dynamics; black: the Italian cumulative curve and in blue the cubic and logistic approximations; green: slow dynamics. b) 7 days scenario. The cubic function (blue-yellow) approximate well the actual cumulative curve (black) also in this scenario, confirming the same result suggested in [27], while the logistic in this scenario performs poorly, and is not in the figure. Note the red dotted curve (12_*_1.342*^n^*), that seems slow, actually is a fast one; instead, the green black dotted curve (16*n*^2^) seems fast, but is very slow.

**Figure 3.**
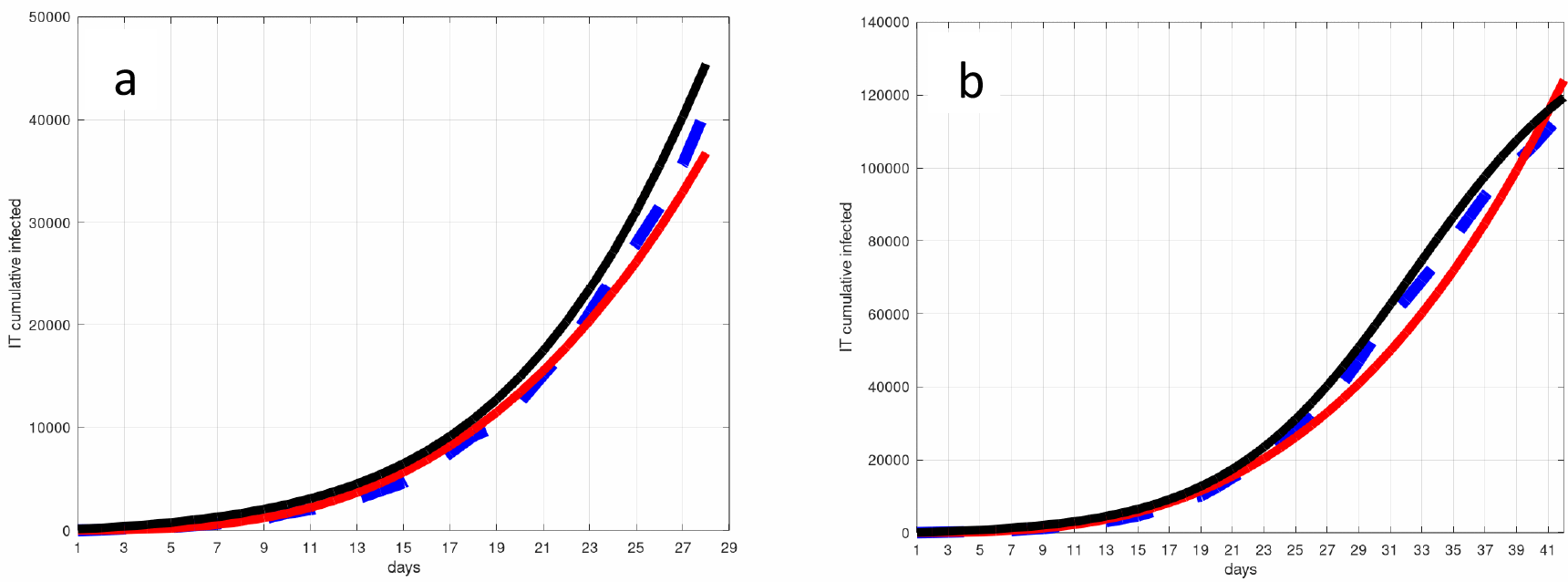
Dotted blue: the Italian cumulative curve, red: cubic approximant, black: logistic approximant. a) first four weeks, the cubic curve is very close to the real growth. b) first six weeks: the logistic curve now is a better approximant.

As said in the above Section, each city provides a sequence of 7 positive integers, and putting them together the fifty sequences make up an unique sequence of 50×7 integers called 50_*cities*. Note that sequences such as {1 2 2 2 2 2 2}, {3 1 1 1 1 1 1}, {1 1 1 1 1 1 1 1}, {1 0.3 … 0.01}, or {16 160 … 16000000}, are not taken into consideration; actually, 8 out of the 50 sequences have been discarded, thus the length of the 50_*cities* sequence is of 294 samples instead of 350.

In the Section above we have indicated that:

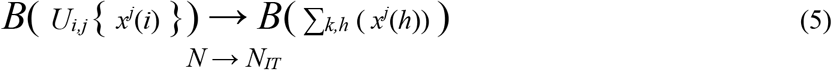

where *i* = 1, 2, … 7. *j* = 1, 2, … 50 *k* = 1, 2, … . *N_IT_ h* = 1, 2, … 42, and *T_f_* = 42 are now specified to the actual case-study scenario.

To classify various possible approximant curves we have calculated their Benford gof, showed in the Table 1, together with the gof of the real Italian epidemic data, the logistic curve, of the cubic curve and of the 50_*cities*.

Keeping in mind that the *union* of the 50 sequences of seven data points from some of the most important Italian cities, converges to the Benford distribution of the *sum* of all the 6 week sequences for all the cities (5), Theorem Berger&Hill 3 [22] guarantee that *almost always* the exponential growth provides numbers according to the NB distribution. Therefore, as explained in other Sections, a good Benford gof obtained from the epidemic cumulative data of the 50_*cities* should be able to identify the type of outbreak dynamic.

In Table 1 the 50 cities gof is 1.8081, very close to the gof for the real Italian data of the first six weeks, *gof_IT* = 1.7313, while the cubic curve gof_*I(t_i_)_*_n*^3^ = 1.8110 has the minimal distance from the 50_*cities* gof. Therefore *n*^3^ is the most probable approximating function, whereas in Figure 3a the Italy’ real epidemic curve is almost coincident with the cubic growth, confirming the results of [27].

Actually, the logistic curve gof is in good agreement with the 50_*cities* gof too, and fits very satisfactorily the Italy’ real data *after* the fourth week (Figure 3b), but the cubic curve has an advantage in terms of Benford gof and till the fourth week is also the best approximant. Therefore, Table 1 reveals that the cubic curve determines the initial stage of the growth.

Again a note of caution: a small gof, say in the interval [0, 3] as in Table 1, is only a *sufficient* condition that guarantees a fast dynamic, but not a necessary one, meaning that a large *gof* > 3 could represent a rapid growth too, as for the case of the function *B*_0_*_*_*6*^n^* . Thus, most of the times, but not always, a large gof indicates a linear or sub-linear dynamics; instead, a small gof guarantees a rapid growth.

Summarizing, the first digit distribution of the 50_*cities* sequence converges to the first digit distribution of the cumulative infected curve, that is Benford during the ascending epidemic phase. If the 50_*cities* gof is small, on the basis of the Berger-Hill’s Theorem we have a fast epidemic growth; instead, if the gof is poor, the growth will be probably slow, although this cannot be assured formally.

Moreover, in order to determine the form of the approximating function that shadows the real epidemic growth, one might extrapolate some of them analytically or heuristically. At this point, it suffices to choose the approximating curve whose gof is close to that of the 50_*cities*. If the difference between the two gof is small (in our case study less than 5%), we assume that the approximating function shadows correctly the real epidemic growth.

## Conclusions

In this paper, we show how to estimate an approximating function of the epidemic curve within a time horizon of five or six weeks, using only the first seven epidemic data points of fifty Italian cities, considered as an unique sequence. The level of compliance of the sequence to the Benford test is used as a criterion to predict the approximating function accuracy with respect to the real epidemic national curve. This procedure is made possible by the convergence of the unique sequence first digit distribution to the Benford distribution, since the national Italian cumulative data first digit distribution is Benford (almost always) when its sequence is a power, exponential or faster curve. Therefore, when a fast epidemic spread is taking place on the territory, its fingerprint will be the Benford distribution, otherwise the spreading will be, with high probability, slow (quasi-linear) or made of sporadic outbursts. Unfortunately, the Benford compliance of the fast growth is only a *sufficient* condition, not a necessary one. Our results make this point clear. On the other hand, early indications about the dynamic nature of the outbreak are made available through a simple statistical test applied to a handful of data. This method has been applied to the Italian covid 19 case study, where the most probable approximating function of the first five weeks has been identified clearly as a cubic curve.

## Data Availability

all data can be freely doenloaded from the link:
https://statistichecoronavirus.it/coronavirus-europa/

## Notes

### Competing Interest Statement

The authors have declared no competing interest.

### Clinical Trial

no clinical trial necessary. only numerical public data have been processed

### Funding Statement

NO funding

### Author Declarations

no details needed

